# Noisy periodicity in tropical respiratory disease dynamics

**DOI:** 10.64898/2026.04.10.26350660

**Authors:** Fuhan Yang, Ephraim Hanks, Jessica Conway, Ottar Bjornstad, Nguyen Thi Le Thanh, Maciej F Boni, Joseph L Servadio

## Abstract

Infectious disease surveillance systems in tropical countries show that respiratory disease incidence generally manifests as year-round activity with weak fluctuations and irregular seasonality. Previously, using a ten-year time series of influenza-like illness (ILI) collected from outpatient clinics in Ho Chi Minh City (HCMC), Vietnam, we found a combination of nonannual and annual signals driving these dynamics, but with unknown mechanisms. In this study, we use seven stochastic dynamical models incorporating humidity, temperature, and school term to investigate plausible mechanisms behind these annual and nonannual incidence trends. We use iterated filtering to fit the models and evaluate the models by comparing how well they replicate the combination of annual and nonannual signals. We find that a model including specific humidity, temperature, and school term best fits our observed data from HCMC and partially reproduces the irregular seasonality. The estimated effects from specific humidity and temperature on transmission are nonlinearly negative but weak. School dismissal is associated with decreased transmission, but also with low magnitude. Under these weak external drivers, we hypothesize that stochasticity makes a strong sub-annual cycle more likely to be observed in ILI disease dynamics. Our study shows a possible mechanism for respiratory disease dynamics in the tropics. When the external drivers are weak, the seasonality of respiratory disease dynamics is prone to the influence of stochasticity.

**Author Summary:** Although the mechanisms driving seasonality of respiratory disease dynamics have been well-studied in temperate regions, they are unknown in the tropics. In this study, we used a 10-year influenza-like-illness (ILI) daily-reporting data set collected from outpatient clinics in Ho Chi Minh City (HCMC) in Vietnam to investigate the mechanisms associated with annual and nonannual (∼215 days) periodic patterns in the data. By comparing seven mechanistic models against the data, we showed that the mechanism that best explains respiratory disease dynamics in HCMC is a stochastic susceptible-infected-recovered-susceptible (SIRS) model weakly driven by external drivers including specific humidity, temperature, and school term. The nonannual cycle’s duration is consistent with the inferred duration of immunity of the model. By showing the nonannual cycle as strong as in the data is only observed in stochastic model, we showed that the observed respiratory disease dynamics in HCMC is under the influence of stochasticity when external drivers are weak.

## 1. Introduction

Seasonality of respiratory disease transmission manifests as regular winter epidemics in temperate parts of the world. The factors associated with this wintertime seasonality have been studied through laboratory experiments [1,2] and modeling studies [3–5]. The common understanding is that climate factors such as humidity [1] and human behaviors such as indoor crowding, movement [6] and school attendance [7–9] are the main drivers of seasonality in temperate regions [10,11]. Significant changes in climate and human activities forced by winter lead to epidemic onset through the increase of viral survival rate and human contact rates when gathering indoors during winter. Globally, the dynamics of respiratory virus transmission are less predictable outside of temperate regions. One obstacle is that there are fewer surveillance systems in subtropical and tropical regions; the lack of data makes it challenging to draw conclusions about seasonality. Another challenge is that seasonality patterns, which are very clear in temperate regions, differ across subtropical and tropical regions. Tropical regions tend to have more irregular seasonality than subtropical regions [12]. Studies have shown that respiratory disease dynamics in subtropical regions present as an annual pattern, a biannual pattern or both [13–15]. As latitude moves close to the equator, dynamics tend to show year-round activity and exhibit weaker fluctuation [14,16,17]. Most studies on respiratory disease dynamics in the tropics are descriptive, focusing on associations between seasonality and environmental drivers [2,13,18–22]. Few studies have examined the mechanisms behind these patterns.

Mechanistically, seasonality of infectious diseases is mainly influenced by four factors: the intrinsic cycle caused host-pathogen interaction [23–25] (like immune waning or pathogen evolution), host behavior, environmental drivers, and stochasticity [26,27]. Seasonality could be influenced by interactions of two or more of these factors, especially, the relative dominance of the interacting factors [28,29]. In respiratory disease dynamics in temperate regions, the regularly annual and substantial change in climate and host behavior may be the dominant factors causing the dynamics to behave annually, overshadowing the influence from host-pathogen interaction or stochasticity [30]. Our previous study found that respiratory disease dynamics in the tropics showed weak fluctuation, with irregular seasonality and both annual and nonannual signals [31]. With no winter in the tropics and mild changes in weather and human behavior, the causes of these fluctuations are not obvious.

We are interested in identifying the mechanisms that cause the unique pattern of respiratory disease dynamics seen in the tropics. In this study, we use seven stochastic dynamical models to investigate the drivers of these dynamics using a 10-year daily respiratory disease reporting dataset from Ho Chi Minh City (HCMC), Vietnam. We incorporate humidity, temperature, and a term-time forcing factor representing school term; all models are based on stochastic versions of a susceptible-infected-recovered-susceptible (SIRS) model. We compare their goodness-of-fit using trajectory matching and their abilities to replicate the cycles observed in the data through power spectrum comparisons. The results of this study show the contributions of environmental factors, school term, and stochasticity to the dynamics, which allow us to identify which of the considered eternal factors can best explain respiratory disease transmission patterns in the tropics.

## 2. Results

We analyze a ten-year standardized daily influenza-like illness (ILI) time series from outpatient community surveillance in HCMC, Vietnam, that has been validated in previous studies [16,31–33] as the main data source in this study. Previous validations on these data included examinations of trends to justify de-trending, evaluation of individual-clinic reporting consistency [33], and evaluation of bias in clinical diagnosis of ILI [31]. Here we scale the time series to represent observed case counts, whereas the previous study presented the time series as a unitless trend in relative ILI incidence. The ILI case time series showed noisy and low-amplitude fluctuations between 2010 and 2019 (Fig 1A). Spectral analysis reveals two periods of comparable importance: signals with cycles of 215 days and 365 days (Fig 1B). This is consistent with our previous results [31], showing that there is a combination of annual and nonannual cycles in the weak fluctuations in ILI dynamics in HCMC.

**Figure 1.**
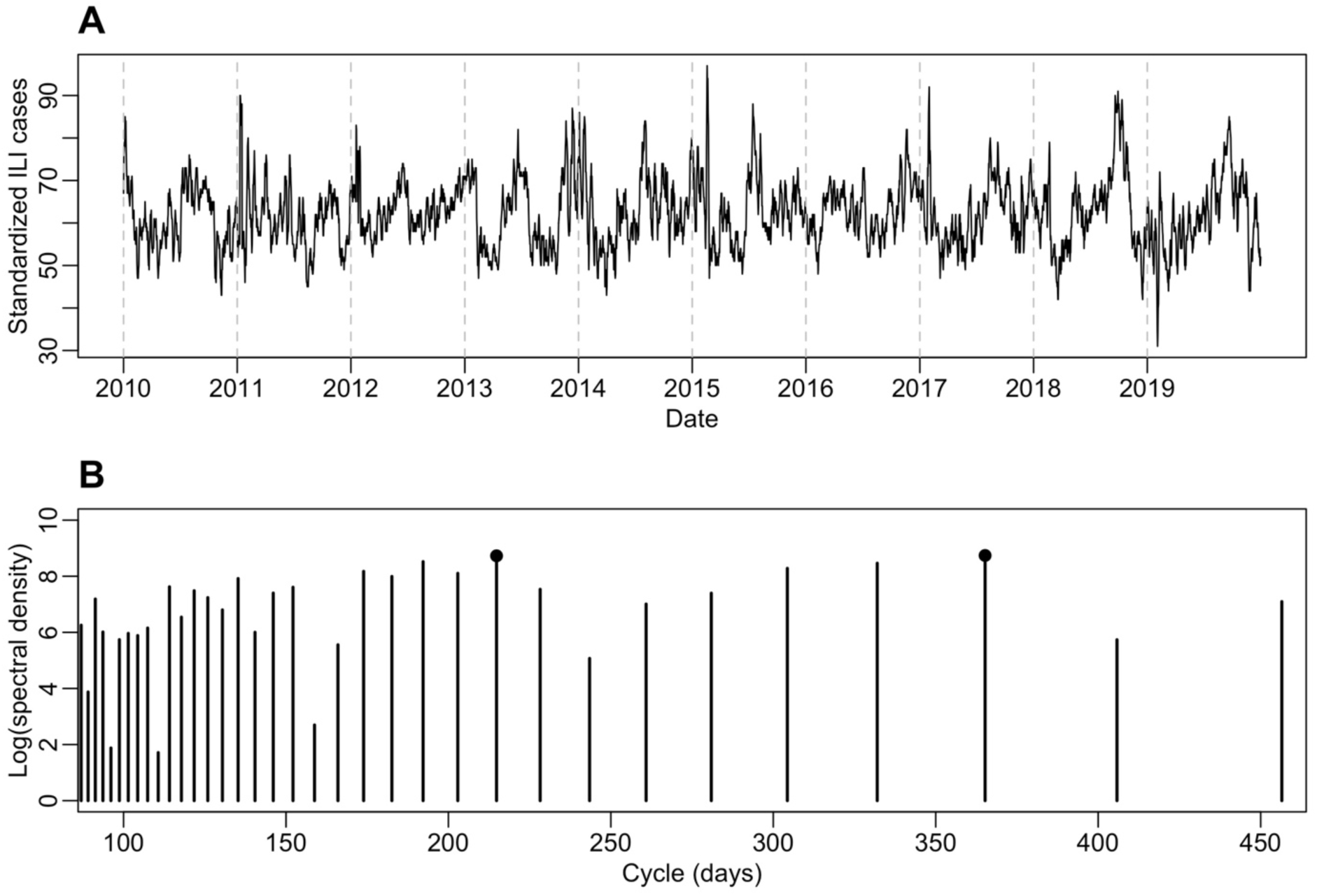
(A) The 7-day smoothed daily ILI incidence from 2010 to 2019 (A). (B) Discrete Fourier transform of the cycles from 100 to 450 days in the ILI data. The two cycles (215 days and 365 days) with the strongest spectral density are labeled with a circle.

### 2.1 Model comparisons through trajectory matching

We describe the respiratory disease dynamics as a model of susceptibles, infecteds and recovereds allowing reinfection (SIRS) using multi-stage infection and recovery classes. We select two I classes (i.e. the infectious period is gamma distributed with shape parameter 2) and three R classes (i.e. duration of immune protection is gamma distributed with shape parameter 3) from a range of possible number of I (1 to 2) and R (3 to 5) classes using partially-observed Markov process framework (SI_2_R_3_S, Fig. S1, details in S2 Text) [34]. We compare the goodness-of-fit of seven models to the data. Respectively, they are: (1) basic SI_2_R_3_S model without external factors (SIRS); (2) SI_2_R_3_S model including absolute humidity covariate (SIRS with AH); (3) SI_2_R_3_S model including smoothly-decreasing school term (SIRS with school term); (4) SI_2_R_3_S model including absolute humidity and school term; (5) SI_2_R_3_S model including quadratic (“U-shaped") effect of specific humidity on transmission, modulated by temperature (SIRS with U-shaped SH and temperature); (6) SI_2_R_3_S model including linear relationship with specific humidity, modulated by temperature (SIRS with linear SH and temperature); (7) SI_2_R_3_S model including quadratic (“U-shaped”) effect of specific humidity, modulated by temperature, and effects of school term (SIRS with U-shaped SH temperature and school term).

The seven models are fitted to the entire ILI time series (January 1, 2010, to December 31, 2019) using iterated filtering [35]. First, we evaluate goodness-of-fit using mean squared error (MSE) between the median from 5000 simulations and the data. SIRS with U-shaped SH, temperature and school term performs the best, indicating that this model best describes the observed data from HCMC. The second-best model is SIRS with U-shaped SH and temperature. The fact that both models share the same feature of the U-shaped effect of specific humidity indicates a level of confidence for this relationship. The average across the 5000 MSE between the data and each simulation is also evaluated to address the variability of individual simulations. With this criterion, the best model is SIRS with U-shaped SH temperature, and the second best adds school term. This means that both models are best able to reproduce the observed data, both across individual simulations and when comparing an aggregate among simulations. In fact, when comparing the median and simulation interval with the data, these two models are the only two models that exhibit small and noisy fluctuations in the data while other models generally produce a visually flat median line (Fig 2). These results suggest the U-shaped effect of specific humidity as a potentially important mechanism of respiratory disease transmission in the tropics.

**Figure 2.**
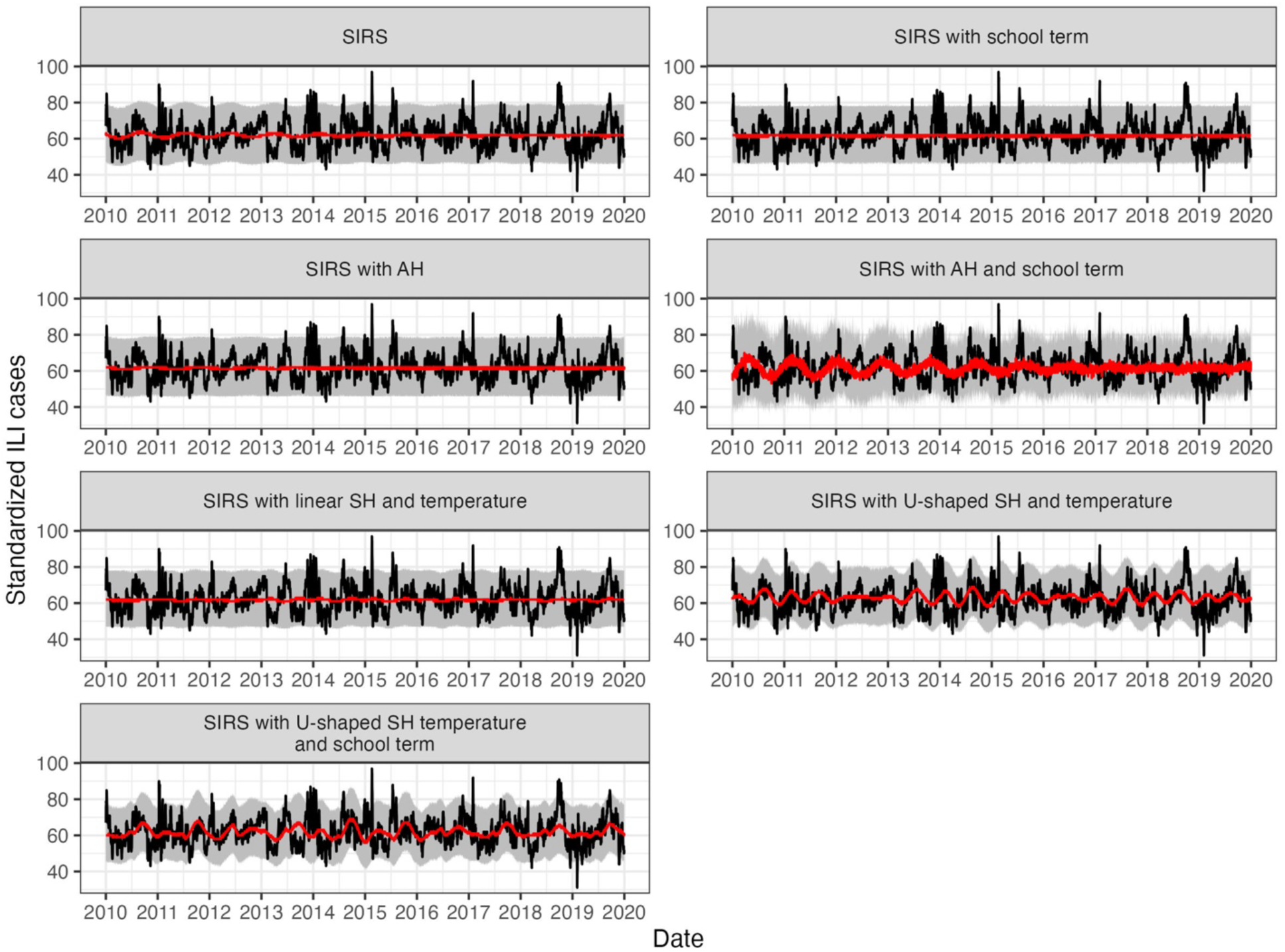
Simulated trajectories from all seven models. The median (red) from 5000 simulated trajectories is plotted with data (black) along with 95% simulation ranges (grey).

Based on the two best supported models, we find that the external drivers have a weak effect on transmission. For SIRS with U-shaped SH and temperature, the parameters governing the U-shaped relationship with SH are estimated to be small in magnitude (*a* = 1.24 × 10^-6^, 95% CI: [1.02 × 10^-6^, 1.29 × 10^-6^], *b* = -1.16 × 10^-5^, 95% CI: [-1.21 × 10^-5^, -1.06 × 10^-5^], Table 1) indicating changes in the transmission parameter *β* many orders of smaller than the estimated value of *β*_0_. Similarly, the modulation effect from temperature is small (*Texp* = 0.088, 95% CI: [0.087,0.090]). These estimations overall show very narrow change in R_0_ (ranging between 1.628 and 1.652) given values of SH and temperature in the entire range observed in HCMC (SH is (9.43 g/kg – 20.65 g/kg) and temperature is (23.11 ℃– 33.31℃)). Similarly, for SIRS with U-shaped SH temperature and school term, the estimated parameters controlling the effect of SH (*a* = 9.36 × 10^-5^, 95% CI: [9.12 × 10^-5^, 9.48 × 10^-5^], *b* = -4.50 × 10^-3^, 95% CI: [-4.58 × 10^-3^, -4.32 × 10^-3^]), temperature (*Texp* = 0.138, 95% CI: [0.136,0.139]) and school term (*a_school* = 4.54 × 10^-3^, 95% CI: [4.47 × 10^-3^, 4.68 × 10^-3^]) on R_0_ are low in magnitude, showing estimated values of R_0_ ranging between 1.550 and 1.602. These results suggest a system near its endemic equilibrium and weakly affected by external drivers.

**Table 1.** Maximum-likelihood estimates (MLEs) of parameters from seven models, estimated R_0_, MSE between the median of 5000 simulations and the data, and the average of 5000 MSE between the data and each simulation. The parentheses show 95% confidence intervals for each MLE, and model rank for two MSE metrics.

| Model/Parameters | SIRS | SIRS with AH | SIRS with school term | SIRS with AH and school term | SIRS with U-shaped SH and temperature | SIRS with U-shaped SH, temperature, and school term | SIRS with linear SH and temperature |
| --- | --- | --- | --- | --- | --- | --- | --- |
| $\beta_0$ | 0.227<br>(0.223,0.229) | 0.213<br>(0.212,0.215) | 0.204<br>(0.196,0.204) | 0.255<br>(0.251, 0.255) | 0.160 (0.160, 0.161) | 0.168<br>(0.167,0.169) | 0.160<br>(0.1600,0.1603) |
| $a_{AH}$ | | -0.55<br>(-0.57,0.53) | | 0.950<br>(0.941,0.951) | | | |
| $w$ | | -99.8<br>(-100, -99.4) | | -23.78<br>(-24.07, -22.71) | | | |
| $a_{school}$ | | | $0.90 \times 10^{-4}$<br>($0.87 \times 10^{-4}$, $0.93 \times 10^{-4}$) | $1.5 \times 10^{-3}$<br>($1.1 \times 10^{-3}$, $1.8 \times 10^{-3}$) | | $4.54 \times 10^{-3}$<br>($4.47 \times 10^{-3}$, $4.68 \times 10^{-3}$) | |
| $\sigma$ | | | 9.69<br>(9.42,9.89) | 4.08<br>(3.37,4.12) | | 19.98 (19.47, 20.00) | |
| $a$ | | | | | $1.24 \times 10^{-6}$<br>($1.02 \times 10^{-6}$, $1.29 \times 10^{-6}$) | $9.36 \times 10^{-5}$<br>($9.12 \times 10^{-5}$, $9.48 \times 10^{-5}$) | |
| $b$ | | | | | $-1.16 \times 10^{-5}$<br>($-1.21 \times 10^{-5}$, $-1.06 \times 10^{-5}$) | $-4.50 \times 10^{-3}$<br>($-4.58 \times 10^{-3}$, $-4.32 \times 10^{-3}$) | $-5.48 \times 10^{-4}$<br>($-6.00 \times 10^{-4}$, $-5.27 \times 10^{-4}$) |
| $c$ | | | | | 0.752 (0.746, 0.760) | 0.70 (0.68,0.71) | 0.50<br>(0.50,0.503) |
| $T_c$ | | | | | 29.27 (29.19, 29.34) | 29.59 (29.32, 29.74) | 29.99<br>(29.81,30.00) |
| $T_{exp}$ | | | | | 0.088<br>(0.087,0.090) | 0.138<br>(0.136,0.139) | 0.0028<br>(0.0026,0.003) |
| $\pi$ | 232.43<br>(231.89, 233.67) | 230.42<br>(227.82, 231.54) | 184.56<br>(183.12, 185.75) | 325.94<br>(325.55,326.63) | 216.89<br>(215.95, 217.79) | 215.78 (215.28, 216.39) | 149.77<br>(148.16, 151.40) |
| $\rho$ | $5.41 \times 10^{-3}$<br>($5.37 \times 10^{-3}$, $5.80 \times 10^{-3}$) | $6.50 \times 10^{-3}$<br>($6.19 \times 10^{-3}$, $6.70 \times 10^{-3}$) | $6.21 \times 10^{-3}$<br>($5.82 \times 10^{-3}$, $6.22 \times 10^{-3}$) | $5.80 \times 10^{-3}$<br>($5.73 \times 10^{-3}$, $5.82 \times 10^{-3}$) | $3.11 \times 10^{-3}$<br>($3.09 \times 10^{-5}$, $3.15 \times 10^{-5}$) | $3.26 \times 10^{-3}$<br>($3.25 \times 10^{-3}$, $3.27 \times 10^{-3}$) | $2.88 \times 10^{-3}$<br>($2.71 \times 10^{-3}$, $3.12 \times 10^{-3}$) |
| $R_{0mean}$ | 1.256<br>(1.232,1.266) | 1.180<br>(1.170,1.190) | 1.124<br>(1.079,1.125) | 1.404<br>(1.382,1.406) | 1.640<br>(1.632,1.647) | 1.575<br>(1.560,1.590) | 1.373 (1.371, 1.378) |
| $R_{0min}$ | | 1.180<br>(1.170,1.190) | 1.123<br>(1.079,1.125<br>) | 1.396<br>(1.376,1.39<br>7) | 1.628<br>(1.620,1.634) | 1.550<br>(1.534,1.565) | 1.371 (1.370,<br>1.377) |
| $R_{0max}$ | | 1.180<br>(1.170,1.190) | 1.124<br>(1.079,1.125<br>) | 1.404<br>(1.382,1.40<br>6) | 1.652<br>(1.644,1.659) | 1.602<br>(1.588,1.616) | 1.378 (1.376,<br>1.382) |
| MSE from<br>the median<br>of<br>simulations<br>(rank) | 63.49 (5) | 63.38 (4) | 68.00 (7) | 63.11 (3) | 61.81 (2) | 60.95 (1) | 64.20 (6) |
| Average<br>MSE from<br>each<br>simulation<br>(rank) | 79.18 (6) | 79.04 (5) | 75.81 (4) | 111.40 (7) | 73.31 (1) | 73.61 (2) | 74.47 (3) |

### 2.2 Model comparison based on power spectrum

To examine a potential cause of the observed annual and nonannual cycle, we compare the power spectrum from 5000 simulations of the fitted models with the power spectrum evaluated from the data. The spectral densities for cycle lengths between 100 and 450 days are calculated for each simulation, and the discrepancies between the data are measured as average weighted MSE (see Methods 5.4).

First, we find that all models with external drivers outperformed the null model (SIRS) in reproducing cycles similar to the data (Table 2). This indicates that the observed cycles are not caused solely by the internal transmission clockwork, but also by external drivers contributing to the dynamics. Second, two SIRS models with U-shaped SH and temperature models (with and without school term) show improved ability in replicating the cyclic pattern of the data compared to other models (Table 2). This suggests that the U-shaped relationship with specific humidity, modulated by temperature, likely contributes to the periodicity of the respiratory disease dynamics in HCMC.

**Table 2.** Average weighted MSE in the power spectrum between 100 days and 450 days across 5000 simulations between each model and the data are shown, with the rank in the parentheses.

| Model | SIRS | SIRS with AH | SIRS with school term | SIRS with AH and school term | SIRS with U-shaped SH and temperature | SIRS with U-shaped SH, temperature and school term | SIRS with linear SH and temperature |
| --- | --- | --- | --- | --- | --- | --- | --- |
| Average weighted MSE across all the simulations (rank) | 11.67 (7) | 10.61 (4) | 11.59 (6) | 9.24 (3) | 6.86 (2) | 5.14 (1) | 11.34 (5) |

We also visualize the power spectrum of the data comparing to the simulations from the seven fitted models (Fig 3). We find that the null model (SIRS) only captures the near-annual cycle (around 330 days). The SIRS with AH, school term, or both, also only captures the near-annual or annual cycles. Although SIRS with linear SH and temperature simulates a bimodal pattern in the cycles same as the data, most of the simulations fail to reproduce the periodic components at nonannual or annual cycles as strongly as observed in the data, indicating that the relationship between specific humidity and respiratory disease dynamics in HCMC may not be linear, which aligns with the results from trajectory matching. Only the models incorporating the U-shaped relationship between SH and transmission modified by temperature (with or without school term) are able to simulate the nonannual cycle at 215 days along with an annual cycle both with comparable amplitudes. This is consistent with the distributions of cyclic summary statistics simulated by the fitted models (S3 Text, Fig S2). Adding school term helps to better reproduce the periodic pattern of the data (Table 2), especially at the annual cycle period (Fig 3). This suggests that school term contributes to amplify the annual cycle, and the nonannual cycle may be the consequences of interaction between climatic factors and respiratory disease dynamics. We also validate the fitting between the best 10 simulations selected by MSE from trajectory matching and the data. We find that these simulations capture the weak fluctuations well (Fig S3). Combining the results from trajectory matching, we select SIRS with U-shaped SH, temperature and school term as the best model capturing the fluctuations as well as reproducing the cycles of respiratory disease dynamics in HCMC.

**Figure 3.**
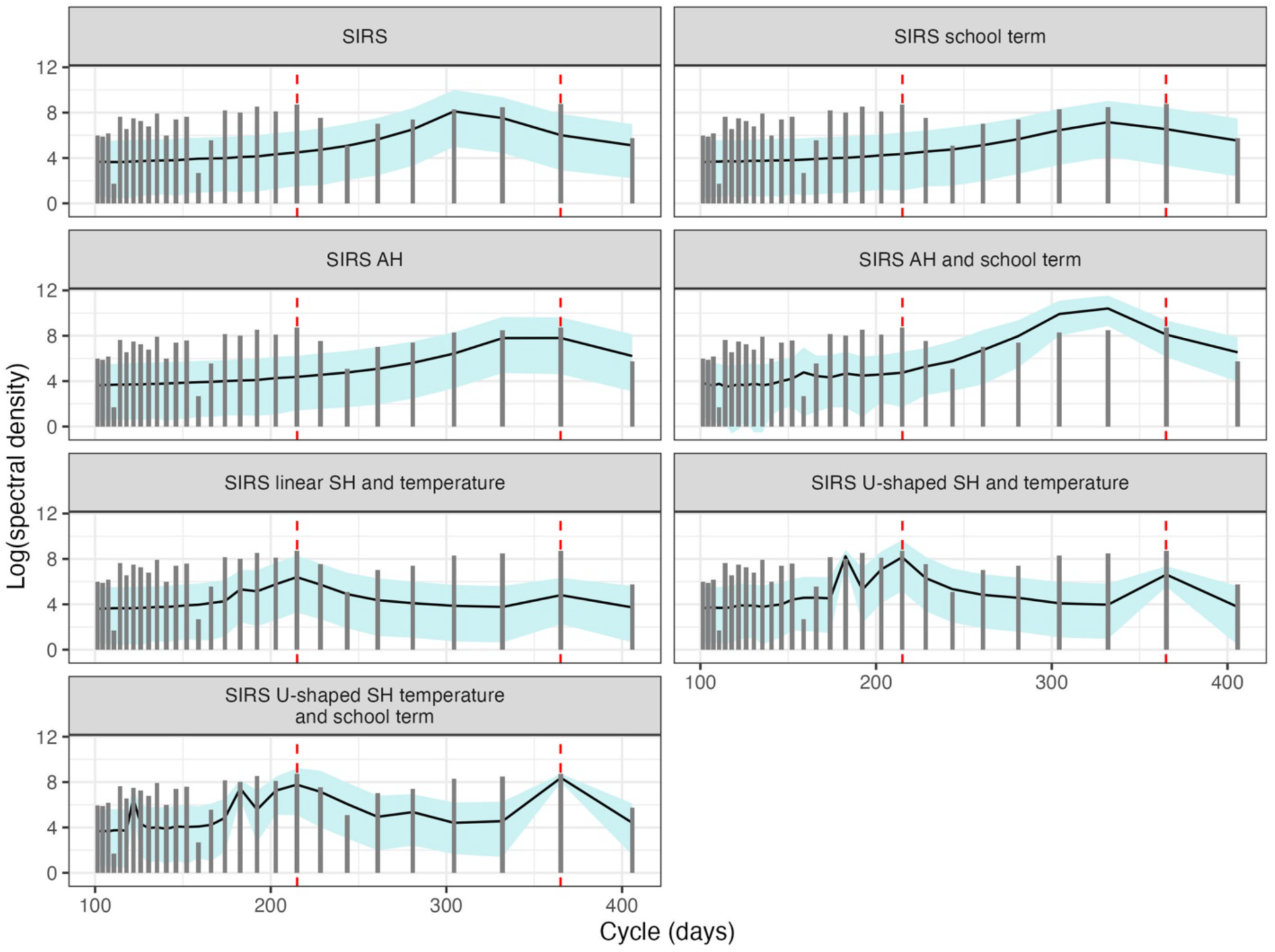
Power spectra from model simulations compared with data. The dark grey vertical bars show the logarithm-transformed spectral density from the data. The mean and the 95% simulation interval of the power spectrum from seven models are shown as black line and blue ribbon. Red dashed vertical lines label cycles of 215 days and 365 days.

### 2.3 Characterization of respiratory disease dynamics in HCMC

We visualize the relationship between specific humidity and estimated R_0_ from SIRS with U-shaped SH, temperature and school term (Fig 4A). We find a weak negative relationship between specific humidity and R_0_. Note that the parameter governing the U-shape of specific humidity is estimated to be very small (*a* = 9.36 × 10^-5^, 95% CI: [9.12 × 10^-5^, 9.48 × 10^-5^], Table 1), leading to a relationship approximated to be negative and nearly linear (*b* = -4.50 × 10^-3^, 95% CI: [-4.58 × 10^-3^, -4.32 × 10^-3^]) as shown in Fig 4A, which means R_0_ will slightly decrease as the SH increases. Similarly, we observe a nonlinear negative relationship between R_0_ and temperature (Fig 4B). As the threshold (*Tc* ) determines whether the effect of temperature is amplifying or inhibiting the effect of SH (see Methods 5.2.e), the estimated high *Tc* (29.59 ℃, 95% CI: [29.32 ℃,29.74 ℃]) suggests temperature will mostly inhibit the effect of SH on R_0_. We can observe that the higher temperature leads to even lower R_0_ (Fig 4A). In addition, we observe R_0_ during school recess is lower than the time of school in session (Fig 4B), while it overlaps with the time when SH is high. However, the overall effect of SH, temperature and school term on R_0_ is weak, leading to a small range of R_0_ over time (1.550 - 1.602).

**Figure 4.**
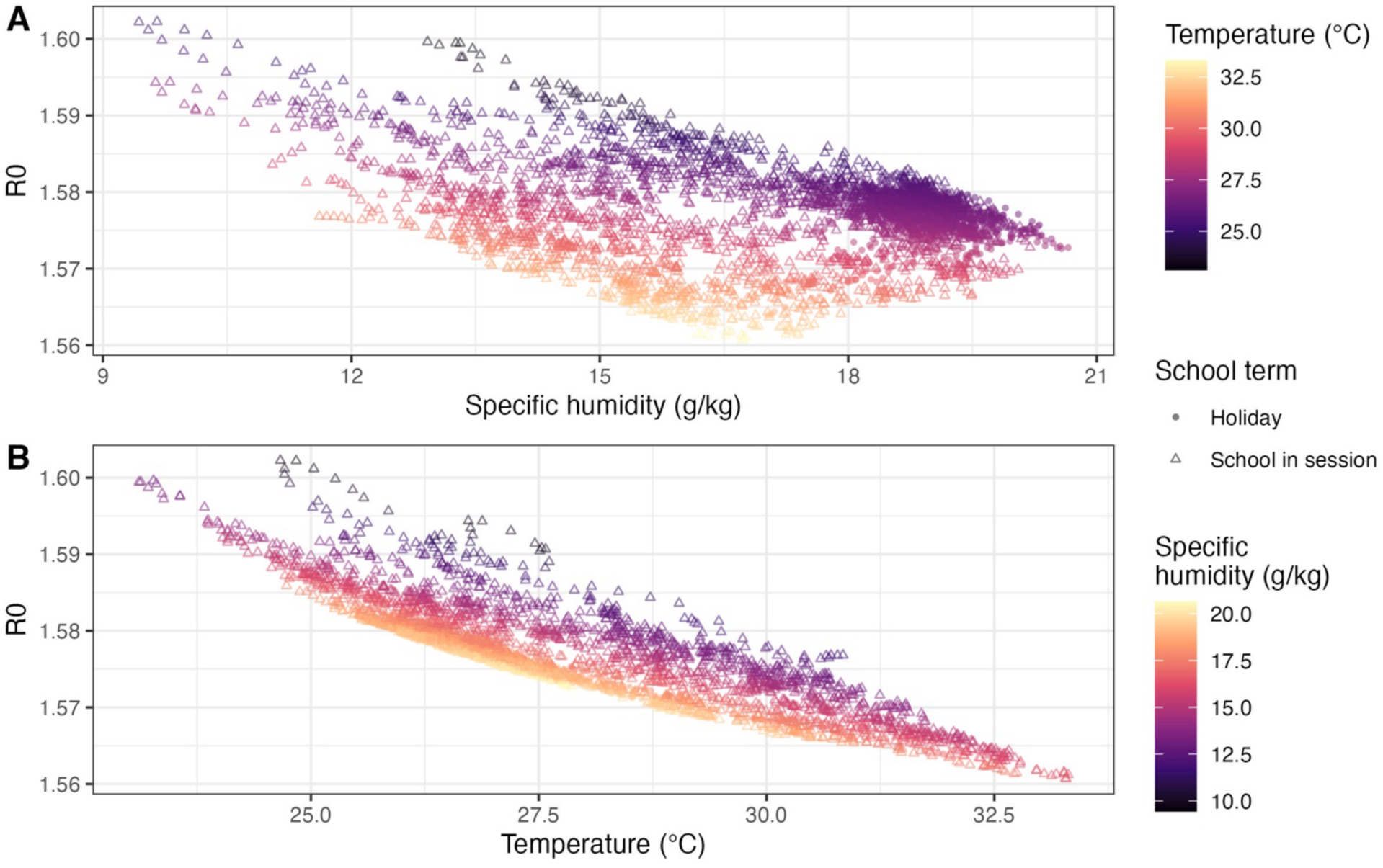
The effect of specific humidity, temperature, and school term on R_0_ from the selected model. (A) The relationship between specific humidity and R_0_ under varying temperatures and school term conditions. (B) The relationship between temperature and R_0_ under varying specific humidity and school term conditions. Triangles show values when school is in session, while circles show schools holidays.

### 2.4 The effects of stochasticity

We compare the simulated power spectra between SIRS with U-shaped SH, temperature and school term and its deterministic skeleton to evaluate the effect of stochasticity on the cycles of the ILI data in HCMC. First, through the average weighted MSE of the power spectra, we find that the power spectrum reproduced by the stochastic model is a slightly better fit than the power spectrum produced by the deterministic version (Table 3), suggesting stochasticity may affect the cycles of respiratory disease dynamics in HCMC. Comparing the power spectrum between data and both models (Fig 5), the annual cycle is well-reproduced by both models, showing the influence from these external annual drivers. However, only the stochastic version is possible to reproduce the nonannual cycle at an amplitude equivalent to the data, or even higher, which the deterministic skeleton failed to do. This indicates that the observed strong signal of the nonannual cycle from the data may be produced under the influence of stochasticity. It is possible that the ILI data in HCMC represents a real-world example that show stochasticity contributes to the seasonality, along with the interaction between external annual drivers and the intrinsic cycle of the disease dynamics.

**Figure 5.**
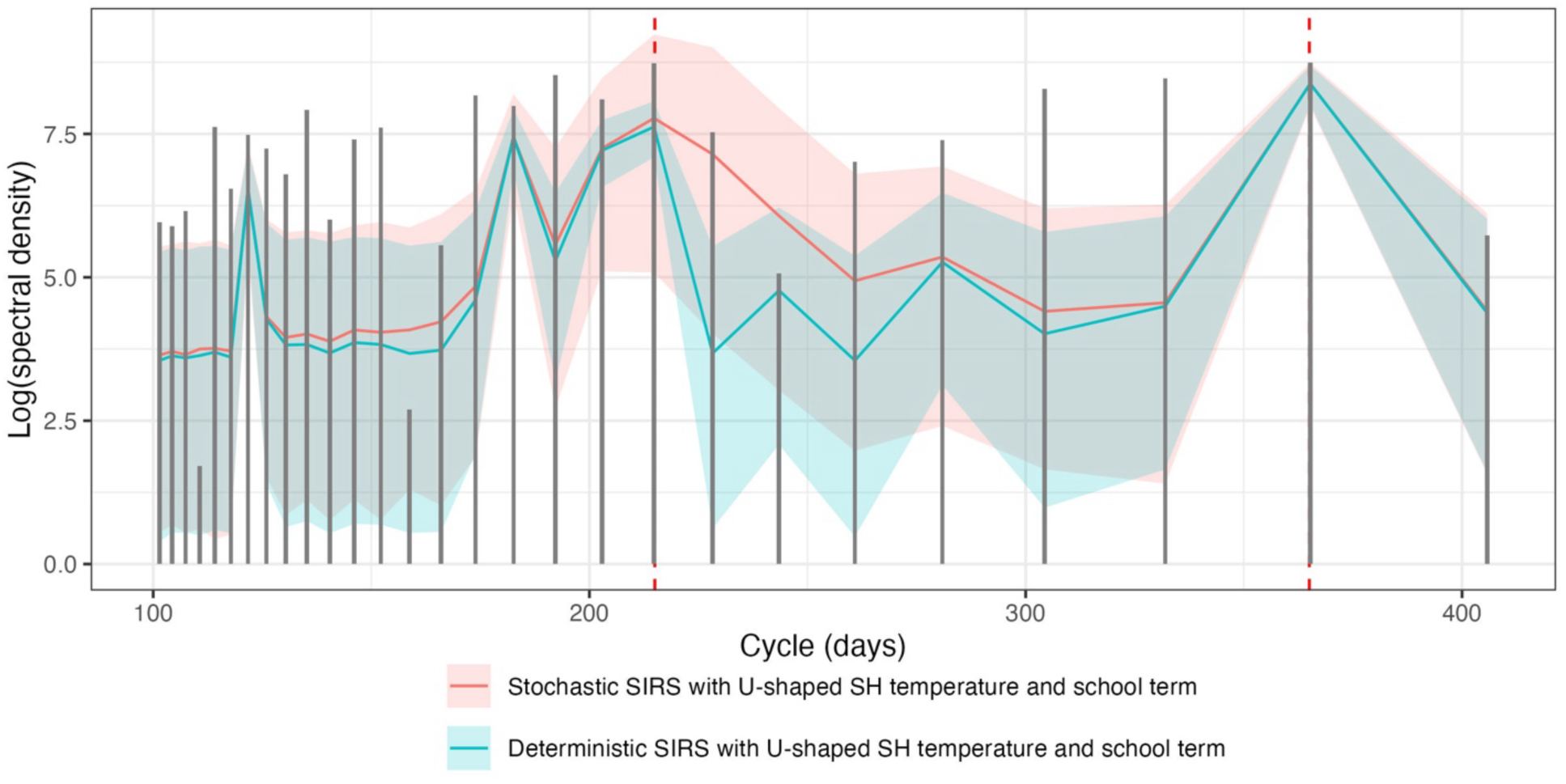
Power spectra of the stochastic SIRS U-shaped SH temperature model with school term and its deterministic skeleton. The power spectrum from the ILI data is shown as grey bars. The mean and the 95% simulation interval of the power spectrum is plotted as red and blue for the stochastic and deterministic version of the model, respectively. The red vertical lines are the cycles at 215 days and 365 days.

**Table 3.** The average MSE in power spectrum from 5000 weighted MSE between the data and each simulation, from stochastic SIRS U-shaped SH temperature and school term and its deterministic skeleton.

| Model | Deterministic SIRS with U-shaped SH temperature and school term | Stochastic SIRS with U-shaped SH temperature and school term |
| --- | --- | --- |
| Average weighted MSE across all the simulations | 5.73 | 5.14 |

## 3. Discussion

We use a 10-year daily ILI time series between 2010 and 2019 to explore the mechanistic forces that drive the unique periodic pattern in respiratory disease dynamics in the tropics, using HCMC, Vietnam, as an example. We compare the performance of seven models on their ability to replicate trajectories and periodicities in the data. Overall, we find that an SIRS model that is nonlinearly influenced by specific humidity, temperature and school term performs best among these seven models. Through this model, we find that specific humidity, temperature and school term drive the transmission negatively but weakly (Fig 4, estimated R_0_ ranges from 1.550 to 1.602 given the entire range of SH, temperature, and school recess in the observed 10-year period). We also find that the observed periodicities are the consequences of interactions between annual external drivers, an intrinsic system cycle (∼215 days), and stochasticity. Specifically, the annual cycle is likely driven by the annual external factors (climate and school term), while the nonannual cycle may be driven by stochasticity. Our study reveals a unique behavior of disease dynamics when external drivers are weak.

This study is one of the first exploring the mechanistic causes of the irregular seasonality of respiratory disease in the tropics [19–21, 39, 40]. There are challenges in studying respiratory disease dynamics in the tropics for two main reasons. First, the absence of high-resolution data over an extended time period makes it difficult to investigate the seasonality of respiratory disease dynamics in the tropics. The ILI surveillance system analyzed here produced more than 66,000 data points from community clinicians over ten years, making it possible to conduct a study of cyclic incidence. Second, it is challenging to provide mechanistic explanations of disease dynamics when dynamical signals are weak. Many descriptive studies have conducted and characterized ILI activity in the tropics as ‘all-year-round’ [14,20,41,42]. Our study is one of the first studies that investigates the lack of regular seasonality of respiratory diseases dynamics in the tropics from a mechanistic perspective.

Fitting noisy incidence data with mechanistic models is challenging. In this case, stochastic models are useful because they can account for noise, which provides flexibility to capture short-term fluctuations that deterministic models cannot catch. Our approach in using stochastic models and stochastic optimization methods allow us to detect a weak dynamical signal behind stochasticity, supporting the utility of stochastic models when fitting noisy data series.

### 3.1 The environmental drivers of respiratory viral infection in the tropics

Our analysis shows that a model with a U-shaped relationship between specific humidity modulated by temperature and school term explains the data the best, consistent with the observations in subtropical regions [37,39]. However, we have not observed the typical U-shape as in previous studies [20,37,38,43], instead observing a monotonic relationship within the observed SH values (the transmission-minimizing SH values were outside the observed range). This may be caused by the estimated effect from specific humidity being weak (*a* = 9.36 × 10^-5^, 95% CI: [9.12 × 10^-5^, 9.48 × 10^-5^], *b* = -4.50 × 10^-3^, 95% CI: [-4.58 × 10^-3^, -4.32 × 10^-3^]). We observe a nonlinear negative effect of temperature on transmission, in alignment with previous observational studies [20] and laboratory findings [44], though this effect is small in magnitude in our analysis (*Texp* = 0.138, 95% CI: [0.136,0.139]). School term-time forcing slightly increases R_0_ (*a_school* = 4.54 × 10^-3^, 95% CI: [4.47 × 10^-3^, 4.68 × 10^-3^]). Overall, our study identifies effects of specific humidity, temperature, and school term on the transmission of respiratory disease dynamics in the tropics, though the effect sizes are small.

### 3.2 Interplay between deterministic forcing and stochasticity

Distinct from respiratory disease dynamics in temperate regions that show a regular annual cycle, the noisy and weak fluctuations with irregular cycles in tropical regions is likely caused by different mechanisms. Our study found that even though the disease dynamics are weakly influenced by the annual cycle of the external drivers, the intrinsic cycle can be observed (∼ 215 days estimated), and stochasticity makes it more likely to see the strong signal of intrinsic cycle (as in the data) (Fig 5). It is possible that the intrinsic cycle of the dynamics and stochasticity dominates when external drivers are weak [28]. The estimated immune-waning duration (π) in our analysis is estimated to be around 216 days for the two best-fit models, suggesting that renewal of susceptibility on this time scale may have an influence on the sub-annual cycle seen in the data. A brute force search for models producing this behavior shows that constructing this effect in a model is non-trivial [45].This kind of the interplay has been widely observed in other infectious diseases like whooping cough [27, 53], rubella [54], measles [53, 55], as well as in the population dynamics in ecology such as flour beetles [56], coastal cod [57], and Dungeness crab [58]. Our results also show that stochasticity could be a cause of the nonannual cycle in HCMC’s ILI activity, suggesting a complicated but remarkable tension in respiratory disease dynamics in the tropics where irregular cyclic upswings would frequently be possible but not always predictable. Perturbation analysis [26] is a natural future direction of this work to more precisely quantify the effect of stochasticity on the cycles in ILI.

### 3.3 Limitations

A major limitation of our study is that our data set includes only symptomatic cases of ILI, which omit asymptomatic infections and include an aggregation of multiple respiratory diseases. However, we have observed consistently weak fluctuation in the ten years of the study period without finding the pronounced regular peak (with a more than 5-fold difference) typically seen in temperate regions [46]. Using our data from HCMC, we also found that ILI cases weakly changed with specific humidity and temperature, suggesting that it is possible that the transmission of the circulating pathogens has an overall weak relationship with environmental drivers in the tropics. While it is also possible that virus interference may attenuate the transmission of a single virus [47], we would require further virological data to understand the transmission of respiratory pathogens in the tropics.

## 4. Conclusions

Our study proposed seven candidate models on the external determinants of the seasonality of respiratory disease dynamics in the tropics. We explored and characterized the assumption that were best able to explain the data through model-fitting as well as power spectrum comparison. We found the dynamics to be weakly driven by specific humidity mediated by temperature as well as by school term. We also found that while these external drivers are weak, the intrinsic sub-annual cycle of respiratory disease dynamics in the tropics may be amplified by stochasticity.

## 5. Methods

### 5.1 Data Acquisition

Between August, 2009 and December 31, 2019, the Oxford University Clinical Research Unit in Vietnam established a community-based mHealth surveillance system to monitor the transmission of respiratory viruses by recording the number of patients presenting with influenza-like-illness (ILI) symptoms in outpatient clinic settings across Ho Chi Minh City [31–33]. A total of 89 clinics participated in this study, and the percentage of ILI patients among the total patients was recorded daily during the period. We used data from 33 clinics for this study because more than 50% of their daily observations report non-zero ILI case counts. Data between January 1, 2010, and December 31, 2019, were used for analysis to remove potential effects of the 2009 H1N1 pandemic. We standardized and smoothed the daily percentage of patients presenting with ILI (details in S1 Text), then multiplied this time series by the 10-year mean of daily ILI cases (62.0 per day) to produce a daily count of ILI cases which we used in this study. The specific humidity and temperature data in HCMC from January 1, 2010 to December 31, 2019 were collected from NASA POWER project [48].

### 5.2 The Models

We use a discrete stochastic epidemic model to model the data, assuming the data is a partial observation of a noisy epidemic process in HCMC. We assume that transmission follows Susceptible – Infected – Recovered – Susceptible (SIRS) dynamics to allow the commonly observed reinfection in respiratory diseases. We incorporate process stochasticity by defining the outflow rate as a binomial draw from each compartment, assuming that the probability is an Euler approximation of the outflow [49].

We include multiple stages in *I* and *R* to model a realistic memory-dependent process in infection and recovery. To select the number of I and R stages, we use a particle filter to evaluate the likelihood of the parameter combinations, including basic transmission rate *β*_0_, reporting rate *ρ*, and immune duration *π*, with each parameter sampled from an educated range (details in S2 Text, Table S1). Two I classes and three R classes are selected as they return the highest likelihood (Fig.S1). The details of Euler-approximated transition rates along with the parameters are listed in Table S2.

Our model can be described as a SI_2_R_3_S epidemic model, which is the model used throughout all the analyses, and we simply abbreviate this as an SIRS model. Seven models are built based on seven variations of the transmission rate *β*, describing seven assumptions about the transmission of respiratory diseases in the tropics. These are listed below.

a. Simple SIRS:

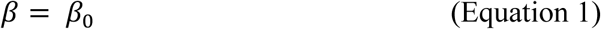 This model assumes constant transmission rate. *β*_0_ will be estimated.
b. SIRS with term-time forcing (SIRS with school term):

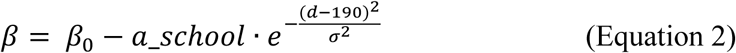 This model assumes the transmission rate is only affected by school term. We assume the transmission rate decreases during the summer vacation, which is from June 1 to August 15 every year. We choose an inverted bell curve (i.e, the shape of an inverted normal distribution) to describe the smooth decrease of transmission during school recess, with the lowest transmission at day 190 of the calendar year, which is July 10, the midpoint of summer vacation. *d* represents the day of the calendar year. In this model, we estimate *β*_0_(baseline transmission, positive), *a*_*school* (peak magnitude of decrease, nonnegative), and *σ* (steepness of bell curve, positive).
c. SIRS with exponential relationship with absolute humidity (SIRS with AH):

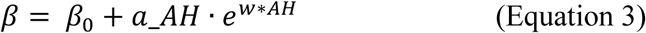 This model assumes the transmission rate is only affected by absolute humidity (AH). The effect of *AH* is transformed through an exponential function and then added to the baseline transmission rate *β*_0_ [1,4]. In this model, *a*_*AH* is an offset term describing the change in the transmission rate when absolute humidity is 0, and *w* is the rate of change of viral viability due to absolute humidity. All of these parameters’ values are estimated. In temperate regions, it is observed that *a*_*AH* is negative, indicating transmission rate decreases monotonically with increasing absolute humidity [3,50]. However, it has not been clear if such relationship exists in the tropics, or if there exists a certain threshold of absolute humidity when switching the relation from negative to positive. Given these reasons, the signs of *a*_*AH* and *w* are not fixed in model fitting.
d. SIRS with exponential relationship with absolute humidity and term-time forcing (SIRS with AH and school term):

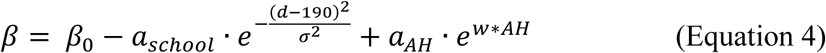 This model assumes the transmission rate is under the composite additive effect of both school term and absolute humidity. The assumptions about signs of parameters remain the same as in the individual-effect models.
e. SIRS with U-shaped relationship with specific humidity modulated by temperature (SIRS with U-shaped SH and temperature):

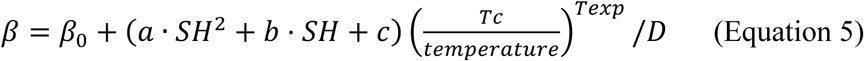 This model assumes the reproduction number *R*_0_ has a non-monotonic relationship with absolute humidity modulated by temperature. It has been observed that the transmission of influenza in subtropical and tropical areas [37,39] possesses this relationship, thus it is used here to test if the transmission in HCMC will follow the same pattern. Specific humidity, a measure of absolute humidity, is applied here. This model assumes the effect of SH on transmission rate is largest when SH is either very low or very high; *a*, *b*, *c* are the coefficients controlling the effect and will be estimated. The effect is moderated by temperature, depending on a threshold temperature *Tc*. When temperature is below *Tc*, the effect of SH on transmission increases and the transmission rate further increases. When temperature is above *Tc*, the effect is decreased, and the transmission is inhibited at higher temperatures. The moderating strength of temperature is controlled by *Text*. Here we estimate the effect of specific humidity and temperature on *R*_0_ on the basis of the baseline transmission rate. We convert the effect to be on transmission rate by dividing the infectious period *D*. The parameters to be estimated are *a*, *b*, *c*, *Tc*, and *Text*. *a*, *b*, *c* are restrained to be positive, negative, and positive, respectively to ensure a minimum value of the parabolic curve and only positive-valued contributions to the transmission parameter. *Tc* and *Text* are constrained to be positive to be valid in describing temperature and its moderating strength.
f. SIRS with U-shaped relationship with specific humidity modulated by temperature and school term (SIRS with U-shaped SH, temperature and school term):

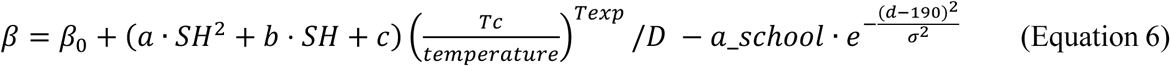 The model assumes additive effect from U-shaped relationship of specific humidity modulated by temperature and school term.
g. SIRS with linear relationship with specific humidity, modulated by temperature (SIRS with linear SH and temperature):

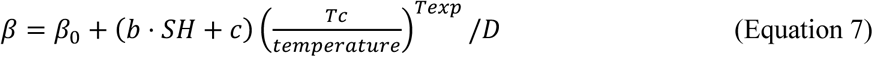 The model assumes a linear relationship between specific humidity and transmission, modulated by temperature. This model is a variant of model e. The parameters to be estimated are *b, c, Tc, Texp. b* and *c* are not constrained. *Tc* and *Text* are constrained to be positive to be valid in describing temperature and the moderating strength.

Reporting rate *ρ* and immune duration *π* of each model are estimated altogether with the parameters from each model. R_0_ is calculated based on: *R*_0_ = *β* × *D*, where *D* represents the duration of the infectious period, which is defined by:

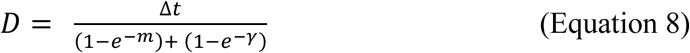

accounting for the Euler-approximated recovery rate (1 – *e*^−*γ*^) and natural death rate (1 – *e*^−*m*^), as well as time discretization, with Δ*t* = 1 in the daily discrete time series. The R_0_ from each model will be calculated with the estimated *β* in this way.

The daily incidence derived from the fitted model is considered as the latent process. We assume the observed data is a Poisson process of the latent process multiplied by the reporting rate *ρ*.

The initial ranges of all the parameters to estimate are in Table S1.

### 5.3 Inference framework

Our model-fitting process uses iterated filtering to maximize a Poisson likelihood function, and we use a particle filter (Sequential Monte Carlo) to evaluate the likelihood. Because of the stochastic nature of the particle filter, likelihood was evaluated 10 times to balance accuracy and computational efficiency given each set of parameters; the standard error of the likelihood is also calculated. We use iterated filtering (IF) algorithm [51] for parameter inference and Monte Carlo likelihood profiling to estimate the 95% confidence interval of the MLEs [52].

Based on preliminary investigations of the likelihood space, we design an inference framework of IF to efficiently search for MLE parameters. We fit the data with four rounds of iterated filtering, conducted sequentially. We first sample 160 sets of parameters from a multivariate uniform distribution for each parameter given educated ranges (Table S1). These sampled parameter sets then undergo iterated filtering four times with different tuning parameters (Table S3). Within the four IF runs, estimates with the 5% highest likelihood are selected as the initial parameters for the next run. The first run quickly explores the entire parameter space and returns the space with a higher likelihood. The subsequent runs conduct more thorough searches as the parameter space is more refined. In this way, the estimates would converge to the global MLE (or close) more efficiently. The likelihood broadly increases along four runs, validating the framework (Fig S4).

The likelihood of the returned parameters from each run is evaluated using a particle filter. The MLE was selected as the run with the highest likelihood and a standard error is smaller than 5. All the analyses used the R package pomp [35] and were implemented in R 4.3.0.

### 5.4 The evaluation of discrepancies in power spectrum

The discrepancies in the power spectrum between the model and the data are measured through average weighted MSE. Firstly, 5000 simulations are generated from each fitted model, and the power spectrum between cycle lengths between 150 and 450 days is calculated for each simulated incidence trajectory. Then, for one simulation, the error for each cycle between the simulation and the data is calculated using MSE. To emphasize how well the model captures the nonannual (215 days) and annual cycles (365 days) in the data, the MSE on these cycles is multiplied by 10 to produce a weighted MSE. Next, the MSE across each cycle is summed to represent the total error in the power spectrum between the data and the simulation. This calculation is repeated for 5000 simulations for each model, and finally the MSE is averaged across 5000 simulations to represent the average discrepancies in the cyclic pattern between models and data, which we call average weighted MSE.

## Data Availability

All the data and code are available at https://github.com/Fuhan-Yang/hcmc_stochastic_ili_modeling.

---

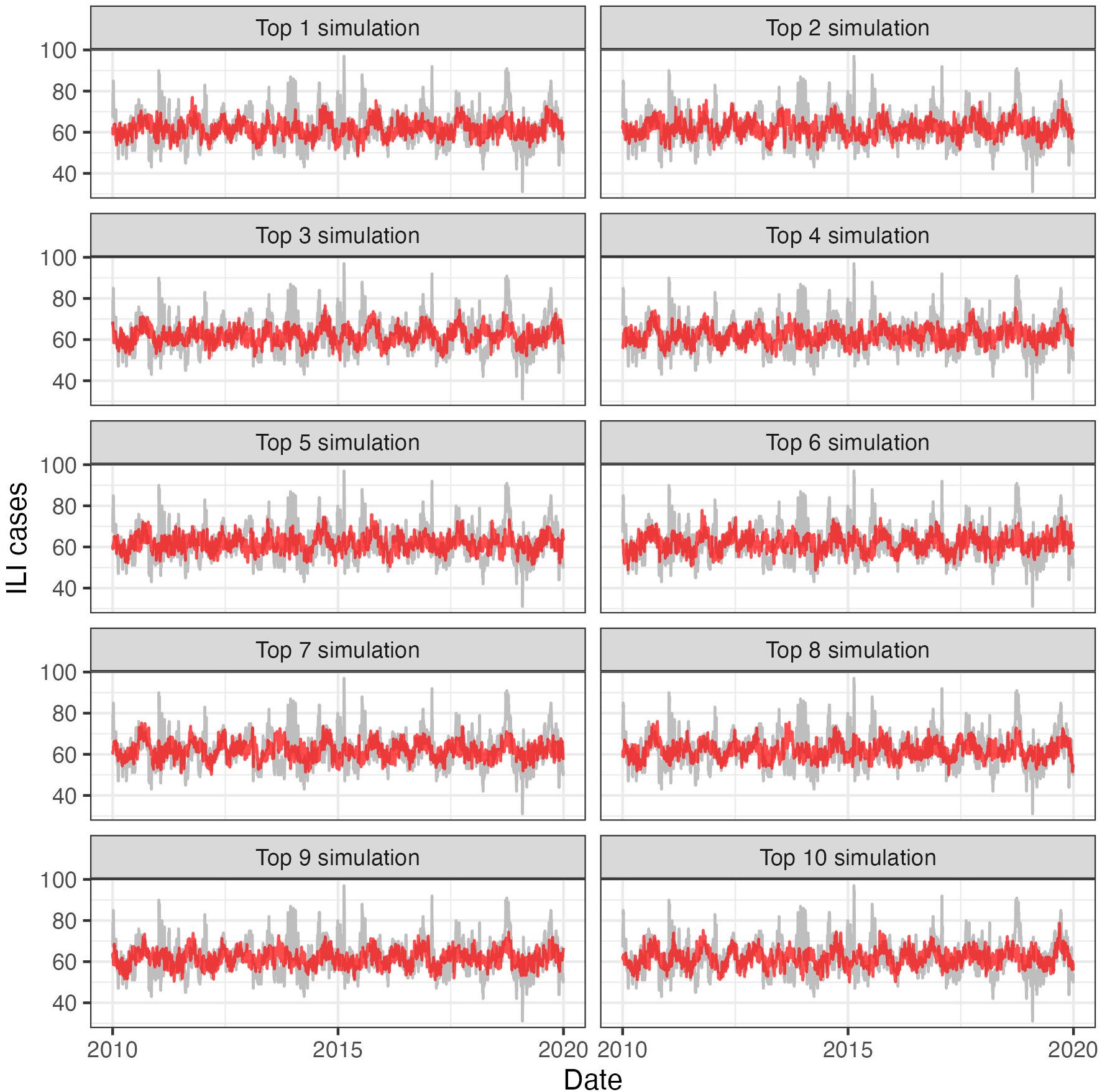

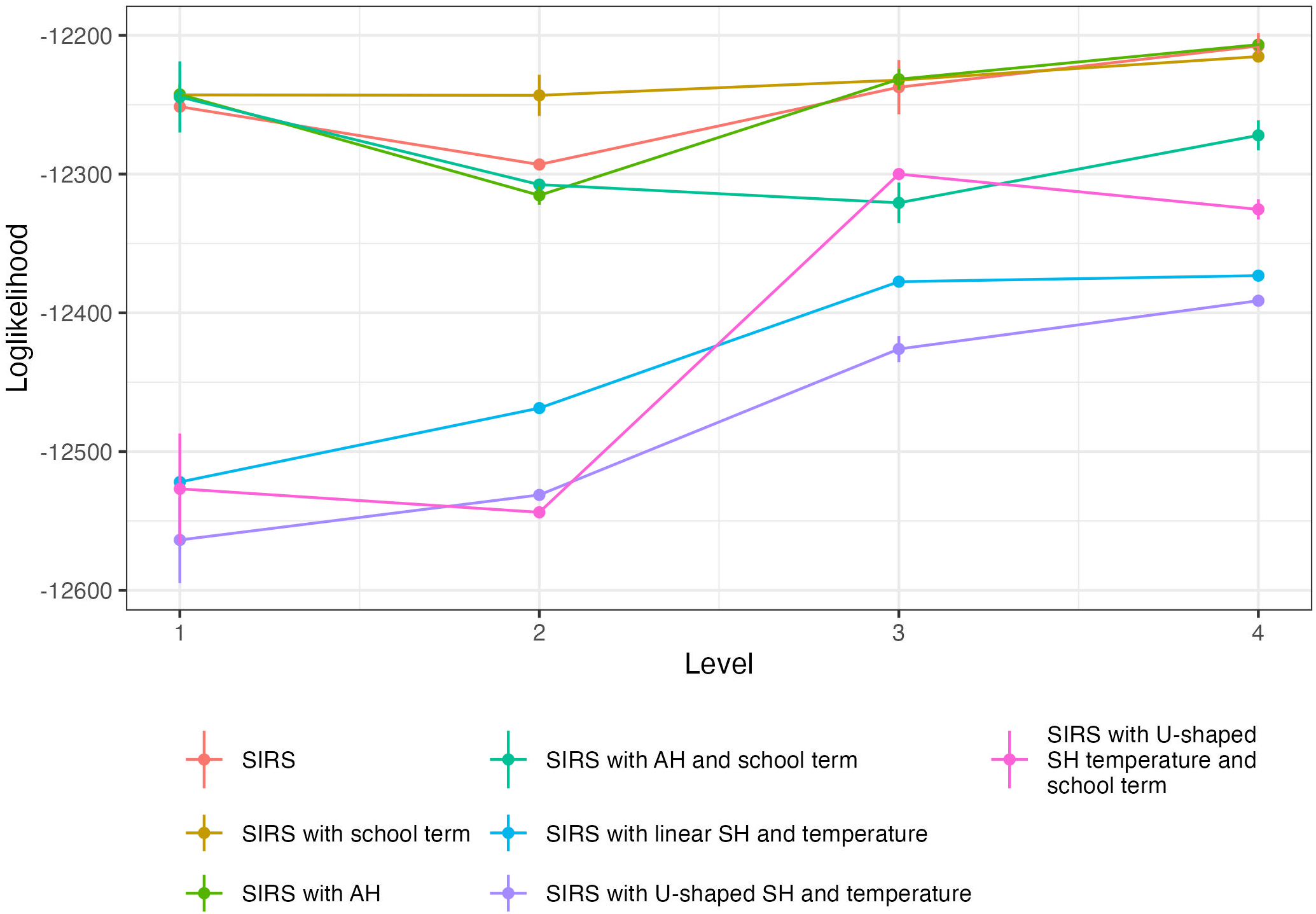

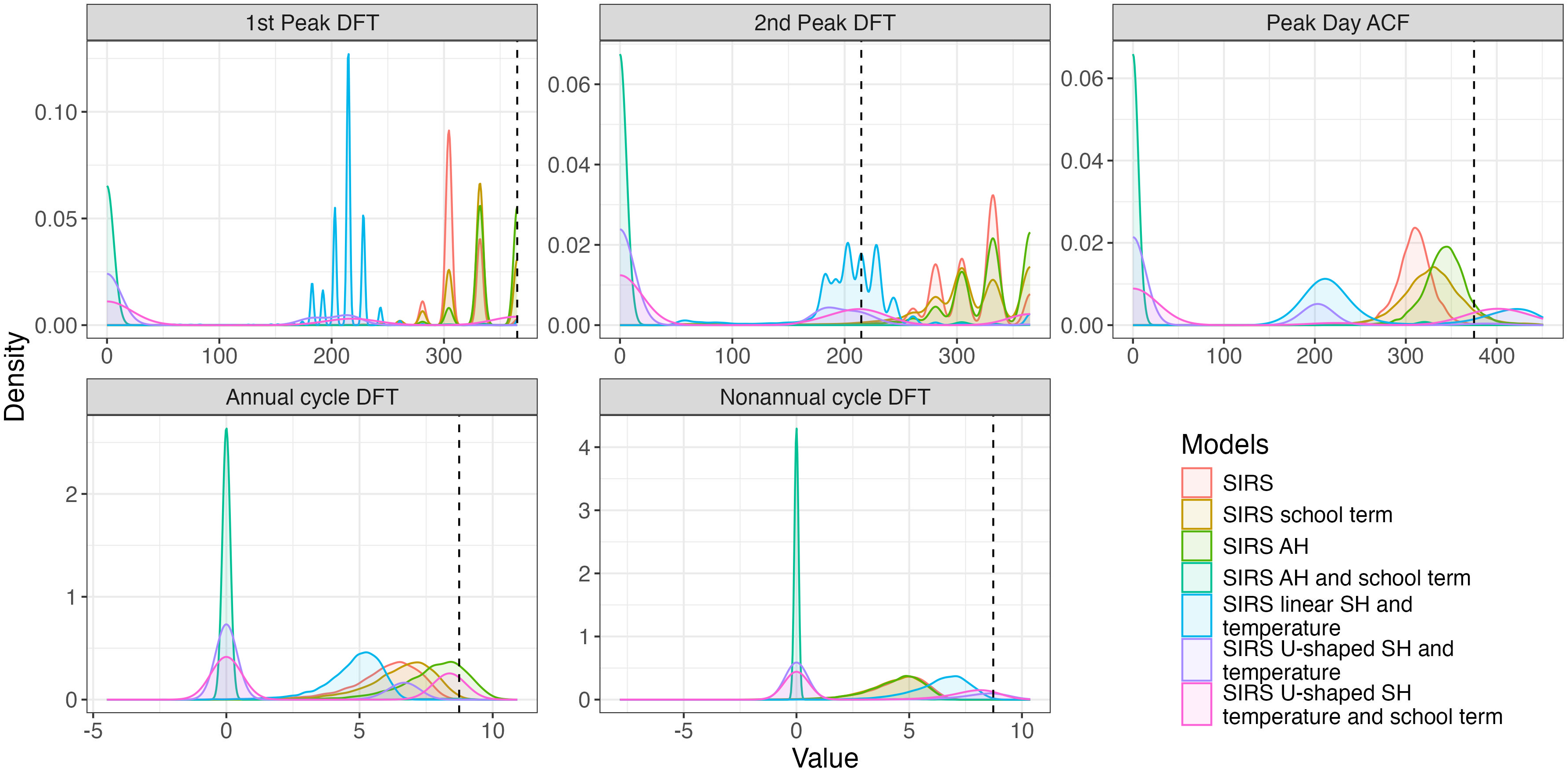

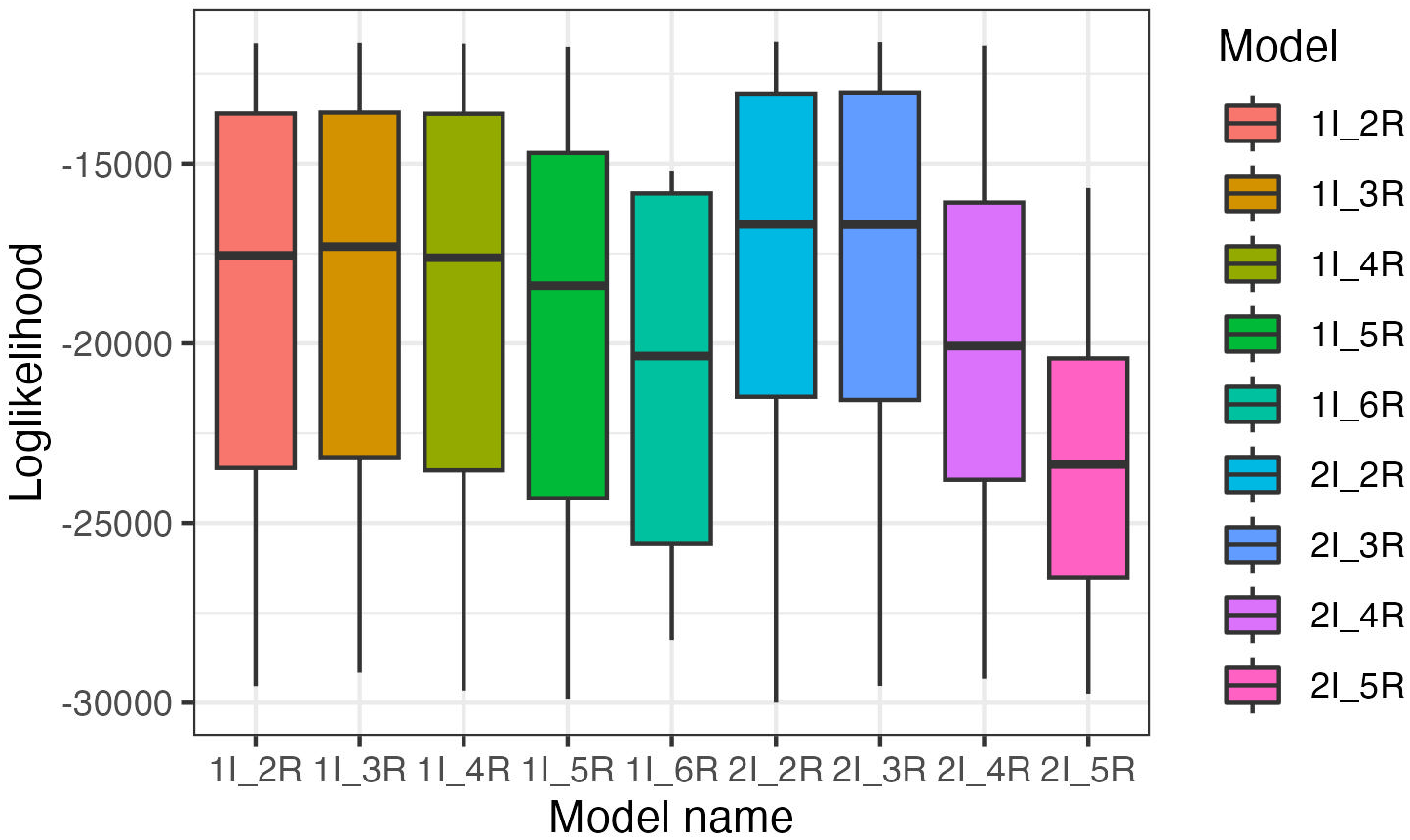

