## Supplemental Materials for "Noisy periodicity in tropical respiratory disease dynamics"

*S1 Text: Time series detrending*

Among the selected 33 clinics, seven clinics show a long-term decreasing trend in %ILI. To remove the trend and get a stationary time series, we detrended the %ILI by dividing each daily value by a 365-day moving average centered at that day for each clinic. We refer to this transformation as a ζ (“zeta”)-score as it is related to an exponentiated z-score. This removes trends longer than 365 days and preserves the ratios between sequential daily reporting numbers. We applied the standardization on the data from 33 clinics and averaged across to get the daily ILI ζ -score. The daily ILI ζ -score was smoothed using 7-day moving average. Then the ILI ζ -score is multiplied by the daily average number of ILI patients observed across the study period (January 1, 2010 to December 31, 2019) and rounded to get the daily ILI cases.

*S2 Text: Choosing the number of I and R stages*

We selected the number of I and R stages in the basic SIRS model (not incorporating external factors) through comparing the likelihood given a range of number of I between 1 and 2, and the number of R between 3 and 5. For each model, the likelihood of parameter combinations composed of $\beta_0$, basic transmission rate, $\rho$,reporting rate, and $\pi$, immune duration is evaluated by particle filter. Each combination includes 5 candidates of each parameter, sampled in uniform distribution given the educated range (Table S1), making 125 parameter combinations for each model. 2000 particles were used to evaluate the likelihood. The model structure with the highest loglikelihood was chosen (Fig S1).

*S3 Text Reproducing cyclic summary statistics*

We performed comparisons of the fitted models to see if they could reproduce periodic patterns of the data, reflected by a series of cyclic summary statistics. Seven models were used to simulate 5000 trajectories from their corresponding best-fit MLE, and the distribution of a series of cyclic summary statistics are plotted, including: (1) the cycles that have the highest spectral density in DFT between 50 days and 450 days (1$^{st}$ Peak in DFT); (2) the cycles that have the second highest spectral density in DFT between 50 days and 450 days were calculated (2$^{nd}$ Peak in DFT); (3) the day lag when the ACF value is the highest between 150 and 450 days (Peak Day ACF); (4) The spectral density at 365-day cycle (Annual cycle DFT); (5) The spectral density at

215-day cycle (Nonannual cycle DFT). We observed that the distribution of 1st Peak in DFT from models with annual drivers (SIRS with school term, SIRS with AH) shows greater density around the observed value in the data, indicating these models are better able to reproduce the strongest signal at annual cycle than other models. However, only the models with U-shaped relationship with SH, adjusted by temperature are able to reproduce both annual and nonannual cycles at comparable amplitudes with the data (Fig S3).

| Model/Parameters | SIRS | SIRS with AH | SIRS with school term | SIRS with AH and school term | SIRS with U-shaped SH and temperature | SIRS with U-shaped SH, temperature and school term | SIRS with linear SH and temperature |
|---|---|---|---|---|---|---|---|
| $\beta_0$ | (0.16,0.4) | (0.16,0.4) | (0.16,0.4) | (0.16,0.4) | (0.16,0.4) | (0.16,0.4) | (0.16,0.4) |
| $\pi$ | (50,500) | (50,500) | (50,500) | (50,500) | (50,500) | (50,500) | (50,500) |
| $\rho$ | (0,0.015) | (0,0.015) | (0,0.015) | (0,0.015) | (0,0.015) | (0,0.015) | (0,0.015) |
| $a\_AH$ | | (-1,1) | | (-1,1) | | | |
| $w$ | | (-100,100) | | (-100,100) | | | |
| $a\_school$ | | | (1e-6,0.1) | (1e-6,0.1) | | (1e-6,0.1) | |
| $\sigma$ | | | (2,20) | (2,20) | | (2,20) | |
| $a$ | | | | | (0,0.5) | (0,0.5) | |
| $b$ | | | | | (-2.5,0) | (-2.5,0) | (-0.1,0.1) |
| $c$ | | | | | (0.5,2) | (0.5,2) | (0.5,2) |
| $Tc$ | | | | | (24,30) | (24,30) | (24,30) |
| $Texp$ | | | | | (0,2) | (0,2) | (0,2) |

Table S1 The initial range of model parameters.

| Rate | Definition | Equation | Parameter |
|---|---|---|---|
| $\mu_{B,t}$ | The daily rate of natural birth, applied to S compartment | $Binom(N, 1 - \exp(-m * dt))$ | $N$: total population in HCMC<br>$m$: daily natural birth count: set as 16 per 1000 per year [2] |
| $\mu_{D,t}$ | The daily rate of natural death, applied to all compartments | $Binom(C_{t-1}, 1 - \exp(-m * dt))$ | $C_{t-1}$,: the number of population corresponding compartment at time *t-1*.<br>$m$: daily natural death count: set as 16 per 1000 per year [2] |
| $\mu_{SI1,t}$ | The daily number of new infections | $Binom\left(S_{t-1}, 1 - \exp\left(-\frac{\beta(I1_{t-1} + I2_{t-1})}{N} * dt\right)\right)$ | $\beta$: transmission rate |
| $\mu_{I1,t}$ | The daily number of infections that enters I2 compartment | $Binom(I1_{t-1}, 1 - \exp(-2\gamma * dt))$ | $\gamma$: the inverse of infectious period, set as 1/5 days$^{-1}$ |
| $\mu_{I2,t}$ | The daily number of infections that enters R1 compartment | $Binom(I2_{t-1}, 1 - \exp(-2\gamma * dt))$ | $\gamma$: the inverse of infectious period, set as 1/5 days$^{-1}$ |
| $\mu_{R1,t}$ | The daily number of people that enters R2 compartment | $Binom(R1_{t-1}, 1 - \exp(-3/\pi * dt))$ | $\pi$: immune duration |
| $\mu_{R2,t}$ | The daily number of people that enters R3 compartment | $Binom(R2_{t-1}, 1 - \exp(-3/\pi * dt))$ | $\pi$: immune duration |
| $\mu_{R3,t}$ | The daily number of people start to lose immunity and enters S compartment | $Binom(R3_{t-1}, 1 - \exp(-3/\pi * dt))$ | $\pi$: immune duration |

Table S2. Definitions of the compartment transition rates in the model given 2 *I* stages and 3 *R* stages along with model parameters. Each rate represents movement between S, I, R states as well as natural birth and death.

| | #iterations | #particles | #iterations after cooling | #particles after cooling | #particles for evaluation |
|---|---|---|---|---|---|
| Run 1 | 20 | 1000 | NA | NA | 2000 |
| Run 2 | 100 | 1000 | NA | NA | 2000 |
| Run 3 | 100 | 1000 | 100 | 2000 | 5000 |
| Run 4 | 100 | 2000 | 100 | 2000 | 5000 |

Table S3 Tuning parameters for each run of the iterated filtering.

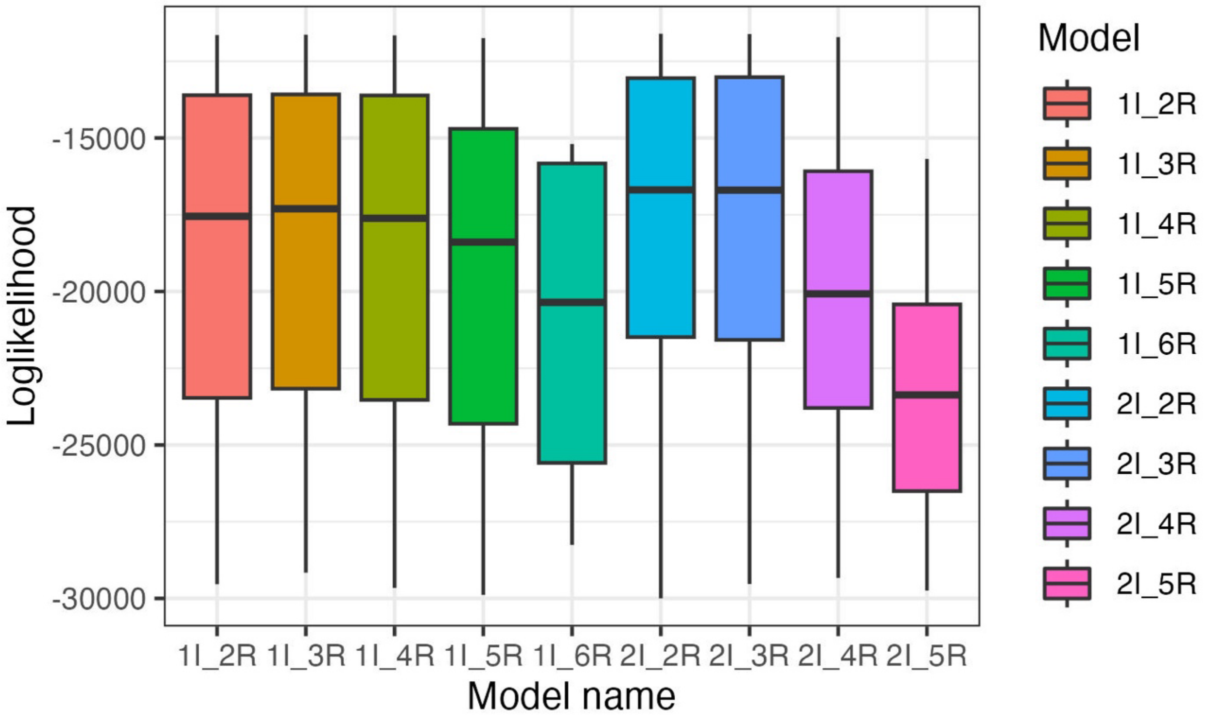


Figure S1. The selection of the number of I classes and R classes. The mean loglikelihood (point) and its standard error (vertical line) of all the estimated parameters was plotted for each model structure. The model with $I_2R_2$ and $I_2R_3$ have similar loglikelihood distribution. The model with $I_2R_3$ returned slightly higher loglikelihood and was chosen for the subsequent analysis.

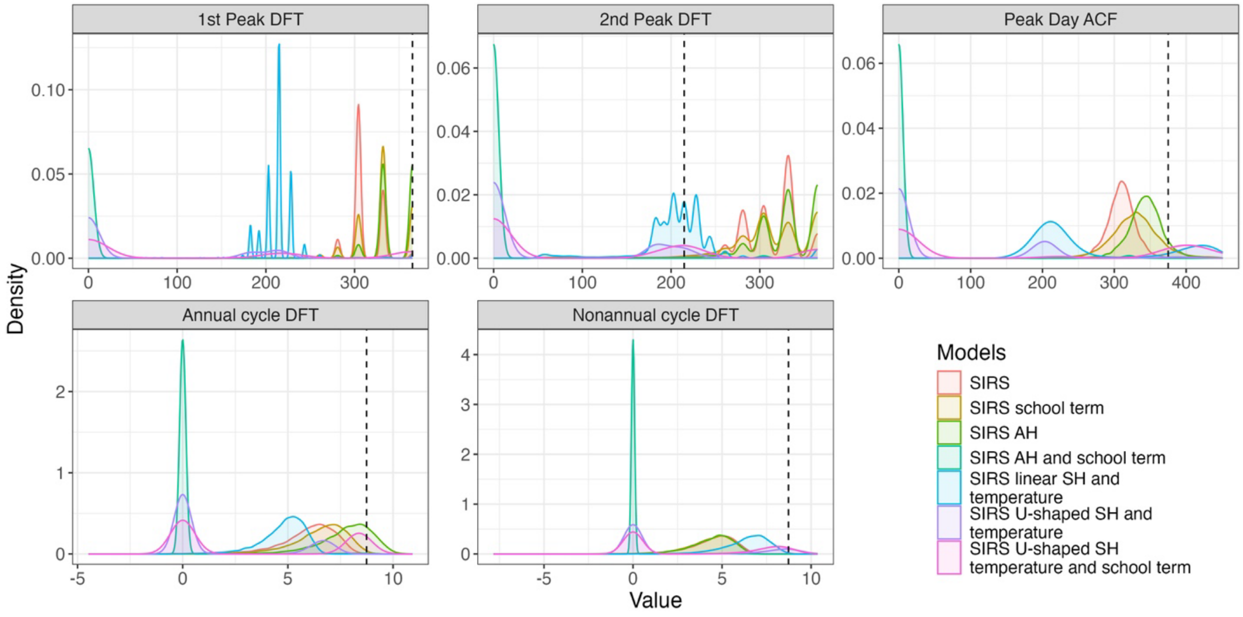


Figure S2 The distributions of the cyclic summary statistics from fitted models. The density of each summary statistics is based on 5000 simulations given the fitted model. The vertical dashed line is the value from the observed data.

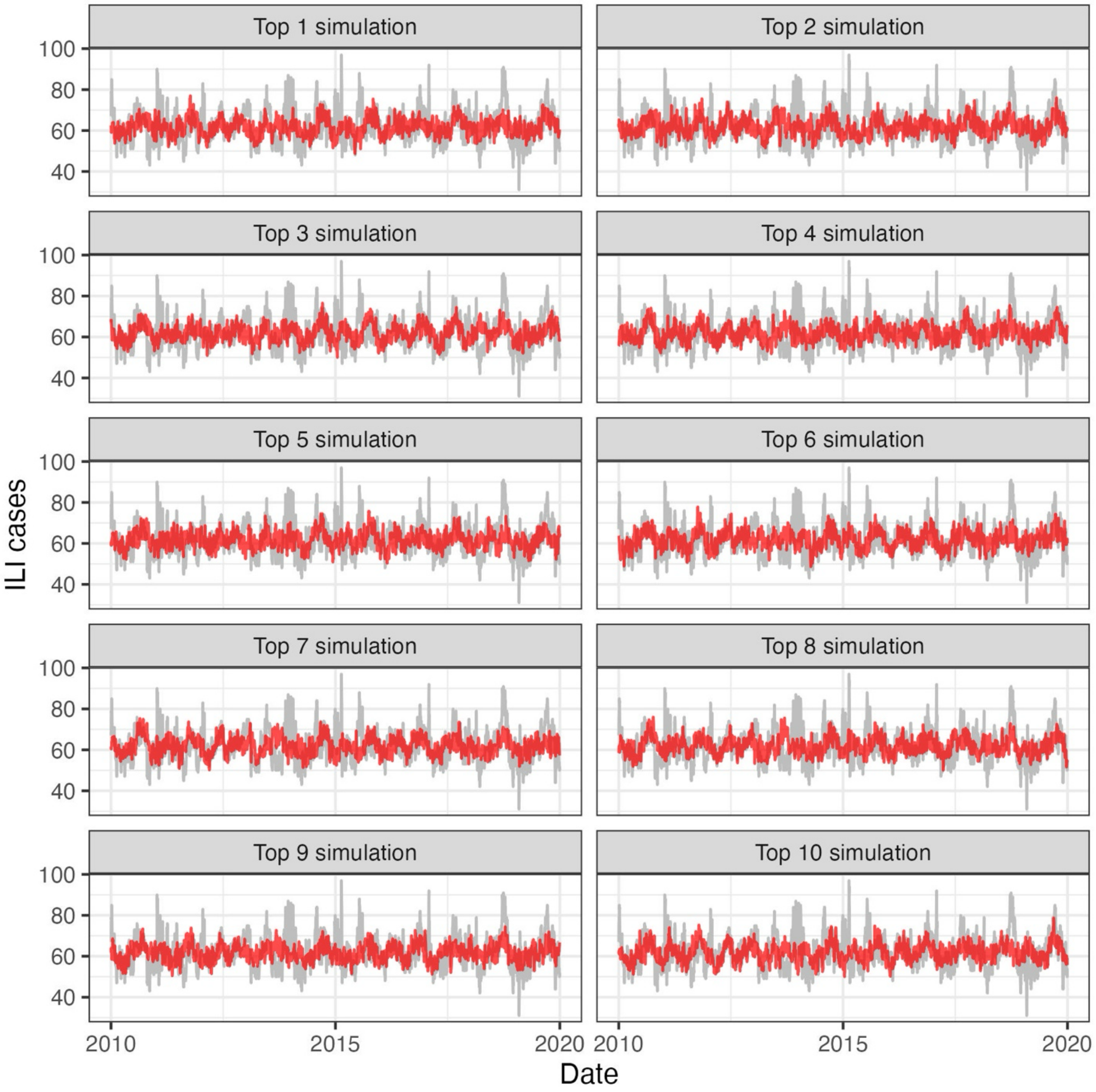


Figure S3 The best 10 simulations chosen from average MSE from trajectory matching of the selected model SIRS with U-shaped SH temperature and school. Simulations are labeled in red; data is labeled in grey.

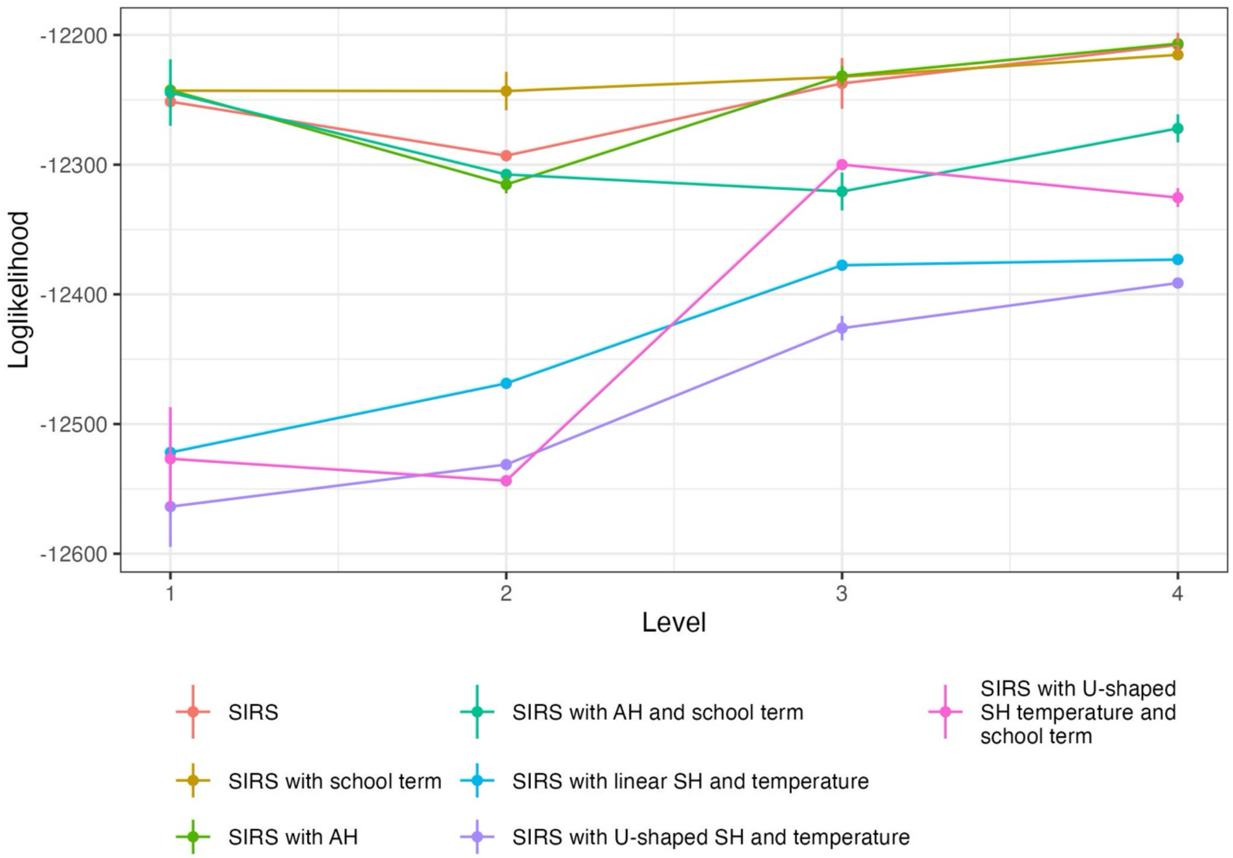


Figure S4. The best likelihood returned for each level of optimization for all seven models. The increase of likelihood from most of the models are observed, indicating the validity of four-run optimization routine. The vertical bar is the standard error of the evaluated likelihood. The occasional decrease of the likelihood may suggest the algorithm is trapped in local optimum on the rugged likelihood surface.
